# Inequities among vulnerable communities during the COVID-19 vaccine rollout

**DOI:** 10.1101/2021.06.15.21258978

**Authors:** Nicholas Stewart, Peter Smittenaar, Staci Sutermaster, Lindsay Coome, Sema Sgaier

**Author notes:** Corresponding author: Sema K. Sgaier.

## Abstract

**Importance:** Federal and state governments sought to prioritize vulnerable communities in the vaccine rollout through various methods of prioritization, and it is necessary to understand whether inequities exist.

**Objective:** To assess whether vulnerable counties have achieved similar rates of coverage to non-vulnerable areas, and how vaccine acceptance varies by vulnerability.

**Design, Setting, and Participants:** We use population-weighted univariate linear regressions to associate the COVID-19 Community Vulnerability Index (CCVI) and its 7 constituent themes with a county-level time series of vaccine coverage and vaccine acceptance. We fit a multilevel model to understand how vulnerability within and across states associates with coverage as of May 8, 2021.

**Main Outcome(s) and Measure(s):** The COVID-19 Community Vulnerability Index was used as a metric for county-level vulnerability. County-level daily COVID-19 vaccination data on both first doses administered and people fully vaccinated from April 3, 2021 through May 8, 2021 were extracted from the Covid Act Now API. County-level daily COVID-19 vaccine acceptance survey data from January 6, 2021 through May 4, 2021 were obtained via the Carnegie Mellon University Delphi Group’s COVIDcast API.

**Results:** Vulnerable counties have consistently lagged less vulnerable counties. As of May 8, the top third of vulnerable counties in the US had fully vaccinated 11.3% fewer people than the bottom third (30.7% vs 34.6% of adult population; linear regression, p= 2.2e-16), and 12.1% fewer initiated vaccinations (40.1% vs 45.6%; linear regression, p= 2.2e-16)). Six out of seven dimensions of vulnerability, including Healthcare System Factors and Socioeconomic Status, predicted lower coverage whereas the Population Density theme associated with higher coverage. Vulnerable counties have also consistently had a slightly lower level of vaccine acceptance, though as of May 4, 2021 this difference was observed to be only 0.7% between low- and high-vulnerability counties (high: 86.1%, low: 85.5%, p=0.027).

**Conclusions and Relevance:** The vaccination gap between vulnerable and non-vulnerable counties is substantial and not readily explained by a difference in acceptance. Vulnerable populations continue to need additional support, and targeted interventions are necessary to achieve similar coverage in vulnerable counties compared to those less vulnerable to COVID-19.

**Key Points:** *Question:* Are the US counties most vulnerable to COVID-19 also facing the lowest vaccination coverage?

*Findings:* US populations with increased health, social, and economic vulnerabilities have experienced consistently lower vaccination coverage. As of May 8, on average, the top third of vulnerable counties across the US had fully vaccinated 11.3% fewer people than the least vulnerable third. There is only a 0.7% difference in vaccine acceptance between the 2 cohorts..

*Meaning:* The gap in vaccination coverage among vulnerable US communities cannot be explained by lower acceptance. Structural barriers need to be addressed to decrease these inequities.

## Introduction

COVID-19 has become the largest global pandemic in over a century. The United States has been disproportionately affected by the virus accounting for 20% of cases and 17% of deaths despite making up only 4% of the population.^1^ Within the US, negative effects spanning health, social, and economic impacts have been disproportionately experienced by the country’s most vulnerable populations. Communities disadvantaged to socioeconomic, epidemiologic, and poorer systemic healthcare settings have taken the brunt of these impacts, and these communities tend to have large minority populations.^2^ Since the start of the pandemic, vulnerable populations have been 20% more likely to have been diagnosed with COVID-19 and 47% more likely to have died.^3^

As we entered the vaccination campaign, disparities in coverage have arisen both at state and local levels.^4^ State- and local-level policies have started to adapt. As of March 31, 2021, there have been 37 jurisdictions, including 34 states, that have used vulnerability indices to prioritize the equitable allocation of vaccines to those most vulnerable.^5^ However, without a universal approach, we will continue to see inequities.

In this study, we sought to analyze if vulnerable communities are continuing to be left behind during the vaccine rollout across the US. We evaluated the association between county-level vulnerability, as defined by the US COVID-19 Community Vulnerability Index^6^, and county-level vaccination coverage. We also analyzed how sentiments of vaccine hesitancy may exacerbate the disparities and if it can explain the gap in vaccination coverage. We hypothesize that vulnerable communities continue to be undervaccinated and that vaccine hesitancy is not the main driver of this disparity.

## Methods

### Vulnerability Index

We define vulnerability specifically to the effects of the COVID-19 pandemic through the use of the US COVID-19 Community Vulnerability Index (CCVI).^6^ As of March 30, 2021, the CCVI was being used by 11 states and 2 cities to prioritize the equitable allocation of vaccines to the most vulnerable populations.^5^ The CCVI is a modular, composite index capturing different dimensions of vulnerability specifically to COVID-19 impacts spanning health, social and economic indicators. The index is a metric, scored from 0 to 1, that compares similar geographic units (e.g., counties) across the US and informs policymakers of communities that require additional support. The CCVI is composed of 7 themes each capturing a different dimension of vulnerability: Socioeconomic Status, Minority Status & Language, Housing Type & Household Composition & Disability, Epidemiological Factors, Healthcare System Factors, High Risk Environments, and Population Density. Overlaying the CCVI with vaccination rates as of June 7, 2021 reveals that the most vulnerable counties also tend to have the lowest vaccination coverage (Figure 1), which concentrates in the South.

**Figure 1:**
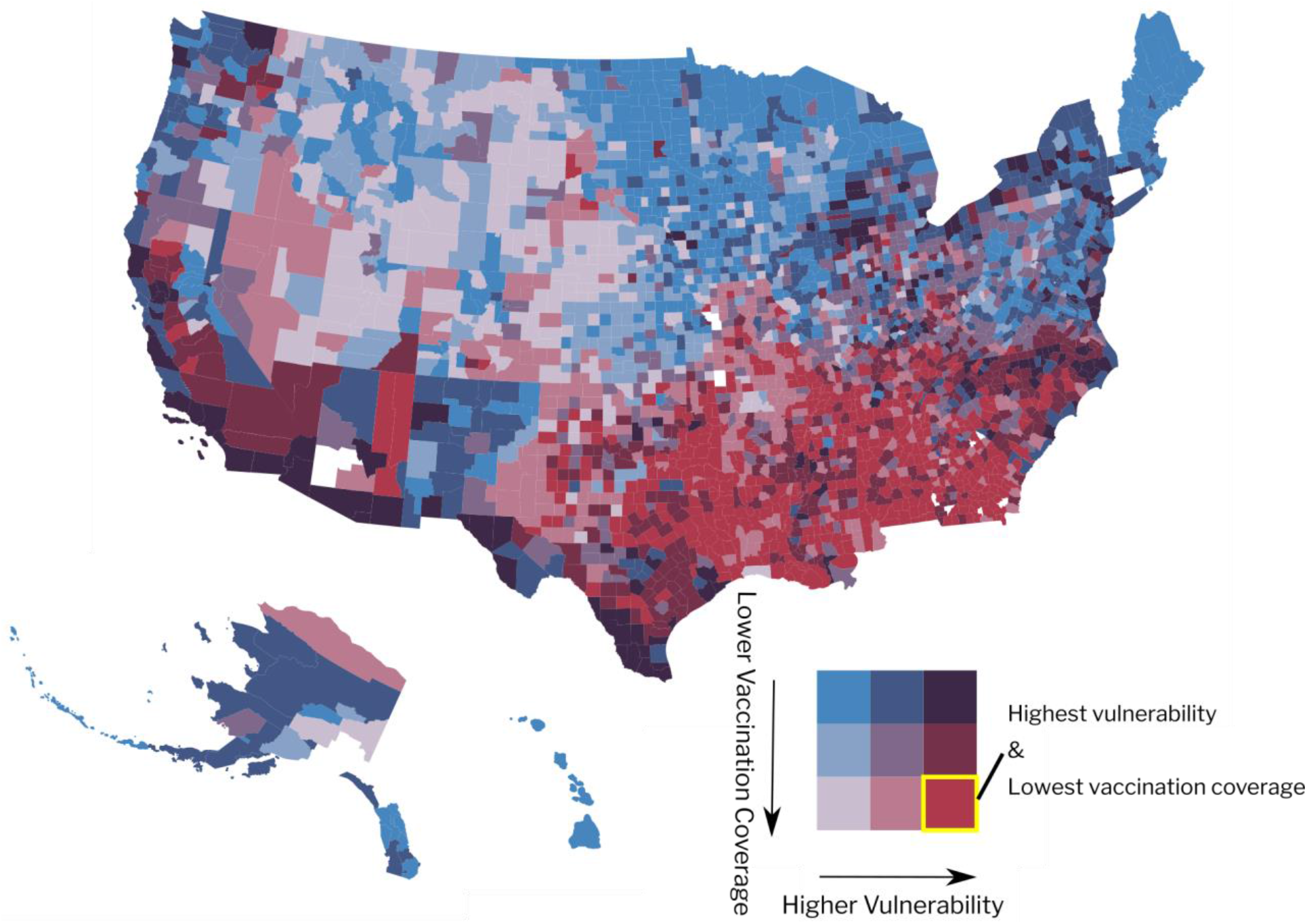
County level bivariate US map shaded by vulnerability (CCVI) and vaccination rates (% fully vaccinated). The vaccination data is from Covid Act Now and is current as of June 7, 2021. There are a total of 3110 counties with vaccination data. The yellow box in the legend highlights the color designating the highest vulnerability and the lowest vaccination rates.

### Longitudinal vaccination rollout data

Daily longitudinal data on the amount of vaccines administered per county were extracted from the Covid Act Now API.^7^ The time series for vaccination data was limited to April 3, 2021 to May 8, 2021 for reasons of data missingness. There are an average of 2,779 counties (out of 3,142) during this time period that make up ∼91% of the total US population with an average CCVI score among the counties of 0.52, which is just above the national average. This ensures that longitudinal analyses conducted on the vaccination data are unbiased and nationally representative. The primary variables of interest include vaccinations initiated per 100 people and vaccinations completed per 100 people as defined by the Covid Act Now.^7^

We obtained daily longitudinal data on county-level vaccine acceptance rates via the Delphi Group of Carnegie Mellon University’s COVIDcast API^8^ from January 6, 2021 through May 4, 2021. Delphi surveys tens of thousands of Facebook users daily regarding their sentiments toward receiving a COVID-19 vaccination. Due to the daily change in both the magnitude and geographic specificity of survey respondents, the introduction of mega-counties is necessary to ensure the data is representative. We assign the daily value of these mega-counties to all counties within. The time series data is representative of an average of 1558 counties comprising ∼61% of the total US population for the given time period. The results of this analysis are therefore biased toward counties with larger populations. The metric of interest is the vaccine acceptance, which indicates the estimated percentage of respondents who *either* have already received a COVID vaccine *or* definitely or probably will choose to get vaccinated if it were offered to them today.

### Statistical modeling of vulnerability against the vaccination rollout

#### Longitudinal univariate regressions

We conducted county population-weighted univariate linear regressions between the vaccination rollout data and the CCVI and its 7 individual themes every day during the study period. We then analyzed the coefficients from those regressions as a time series to understand the association between vaccination coverage and vulnerability as it evolved throughout April and early May 2021. We also analyzed the R^2^ values to understand the variance in coverage explained by the CCVI and its 7 themes. To more deeply understand the association between minorities & language vulnerability and coverage, we performed the same regressions but using the individual indicators (percentage population minority and English as a second language, respectively).

The same county population-weighted average univariate regression models were performed using vaccine acceptance data to understand the association between populations that are vaccine hesitant and their vulnerability characteristics. The model was again applied to the individual variables of percentage minority and percentage of limited English speakers to evaluate the relationship between minority populations and negative sentiments toward receiving a vaccine.

#### Multilevel Modeling

Vaccination coverage might be associated with vulnerability at two levels. Firstly, within a state, more vulnerable counties might be slower to vaccinate relative to less vulnerable counties within the same state. Secondly, more vulnerable states might be slower to vaccinate than less vulnerable states. Disentangling which associations exist - intrastate, interstate, or both - has implications for what interventions are needed to support vulnerable communities. We conducted mixed effects modeling with a fixed effect for the deviation of the county’s vulnerability from its state vulnerability (intrastate variation), one fixed effect for the county’s state vulnerability relative to all other states (interstate vulnerability), and a random intercept for states.^9,10^

## Results

As of May 8, 2021, counties across the US had administered, on average, the first dose of the COVID-19 vaccine to 41.4% of their population and fully vaccinated 31.7% (among the 2968 counties with available data). The top third of vulnerable counties administered first doses to only 40.1% and fully vaccinated 30.7% of their population. By comparison, the bottom third of vulnerable counties administered first doses to 45.6% and fully vaccinated 34.6% of their population, i.e., 14% and 13% more, respectively. The same comparison by each of the seven themes shows more vulnerable regions have fully vaccinated fewer people, except for the minority & language theme and population density theme, for which more vulnerable counties have achieved greater coverage (eTable 1 in the Supplement).

### Variation in vaccination coverage by vulnerability types

Six out of the 7 vulnerability themes were significantly associated with lower vaccination coverage during April and early May (Figure 2). The strongest associations were seen for counties scoring high on the CCVI and for three themes: Housing Type, Transportation & Household composition; Healthcare System factors; and Socioeconomic Status, across both first doses administered and percentage fully vaccinated.

**Figure 2.**
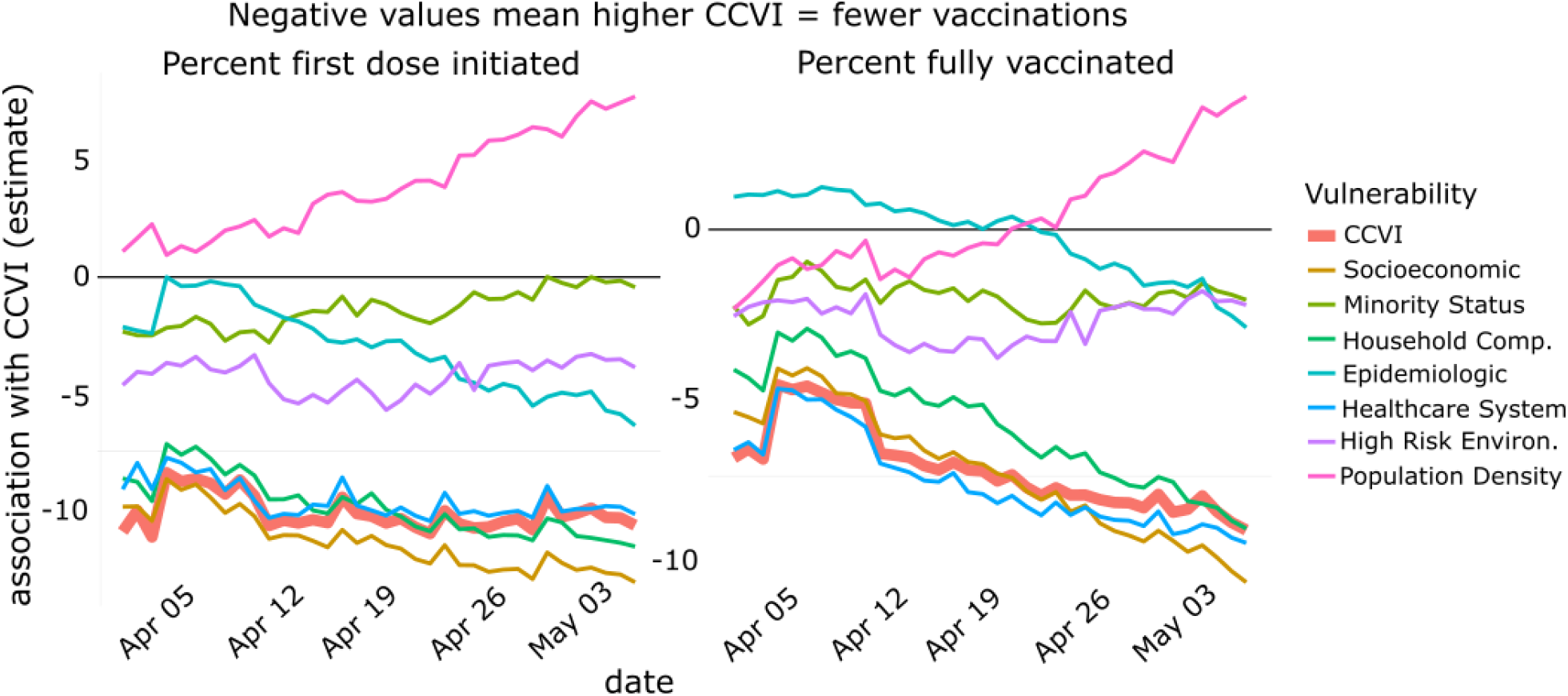
Regressions are simple univariate linear regressions, no mixed effects, weighted by county population. The dependent variable is the cumulative percentage of the total population that have received at least 1 dose (A) and that have completed vaccination (B). The coefficient of variance from each daily regression is plotted as a time series.

The relationship between higher CCVI and those three theme scores with lower vaccination coverage grew stronger throughout the month of April, further increasing the gap in coverage. Prior to mid-April, counties with larger populations with Epidemiologic risk factors tended to have greater vaccination coverage, however this association then reversed. Not all vulnerability dimensions were associated with decreases in vaccination coverage. In mid-April 2021, denser urban environments transitioned from a negative to a positive correlation with vaccine coverage compared to more rural geographies. This trend continued to strengthen into May. The top third of counties with the highest population density have seen 18.8% more fully vaccinated people as of May 8. Minority Status & Language vulnerabilities, Epidemiologic factors, and High Risk Environment factors all had weaker associations but tended to correlate negatively with vaccine coverage.

### Disentangling vulnerability across and within states

The results from the multilevel analysis (Table 1) indicated that vulnerability was associated with vaccination coverage both among counties within-state and across states. The intercept for the model regressing against the overall CCVI was 27.6 indicating that for a county with an average CCVI score, 27.6% of the population had been fully vaccinated. On average, the counties in the most vulnerable states had vaccinated 9.4 percentage points fewer people than the least vulnerable states (Table 1, column 3; p=.004). The effect of the CCVI within a state was less severe, but still significant. On average, for every unit increase in the CCVI for counties within the same state, the fully vaccinated population decreased by 5.4 percentage points.

**Table 1.** Results from multilevel regression models between centered vulnerability metrics (i.e., CCVI and its 7 themes) and vaccination coverage (i.e., percent fully vaccinated per county).^1^

| Independent variable | Intercept | CCVI across states (p-value) | CCVI within state (p-value) |
| --- | --- | --- | --- |
| CCVI (overall index) | 27.73 | -9.43 (**0.004**) | -5.4 (**<0.001**) |
| Socioeconomic Status | 28.04 | -9.14 (**0.017**) | -6.44 (**<0.001**) |
| Minority Status | 28.36 | -2.23 (0.561) | 0.92 (0.094) |
| Housing Type/HH Comp | 28.22 | -6.27 (0.208) | -5.82 (**<0.001**) |
| Epidemiological Factors | 28.09 | -4.23 (0.257) | 1.92 (**<0.001**) |
| Healthcare System | 27.52 | -8.47 (**0.003**) | -11.79 (**<0.001**) |
| High Risk Environments | 27.66 | -11.08 (**0.010**) | 0.13 (0.78) |
| Population Density | 28.3 | 1.31 (0.710) | 3.19 (**<0.001**) |

Only 2 themes, Socioeconomic Status and Healthcare System Factors, were associated with lower vaccination rates among counties both across and within states. Higher Housing Type & Household Composition vulnerability, lower Epidemiologic Risk Factors, and lower Population Density correlated with lower vaccination rates among counties within state only. However, higher scores of High Risk Environments were associated with lower vaccination coverage among counties across states only.

### The CCVI in the context of vaccine hesitancy

As of May 4, 2021, the average county-level population of vaccine acceptors across the US was 85.9% (among the 1558 counties with available data). We split these counties into terciles according to the CCVI and found that the top third of vulnerable counties had a population of vaccine acceptance of 85.5% (values for all other themes can be found in eTable 2 in the Supplement). By comparison, the bottom third of vulnerable counties had a population of vaccine acceptance of 86.1%, a mere 0.7% difference (p = 0.027).

We plotted a time series of correlation coefficients between the CCVI and vaccine acceptance and found that the CCVI and 5 out of the 7 themes were significantly associated with lower vaccine acceptance during the month of April and early May (Figure 3). The strength of the relationship between the vulnerability metrics and vaccine acceptance remained mostly unchanged throughout the study period. A unit change in the overall CCVI resulted in an average 5% decrease in the population of vaccine acceptors during the study period. The Epidemiological theme had the strongest correlation with lower vaccine acceptance. Not all vulnerability dimensions correlated with lower vaccine acceptance. Denser urban environments (i.e., Population Density) and minority populations tended to have more vaccine acceptors throughout April and early May.

**Figure 3.**
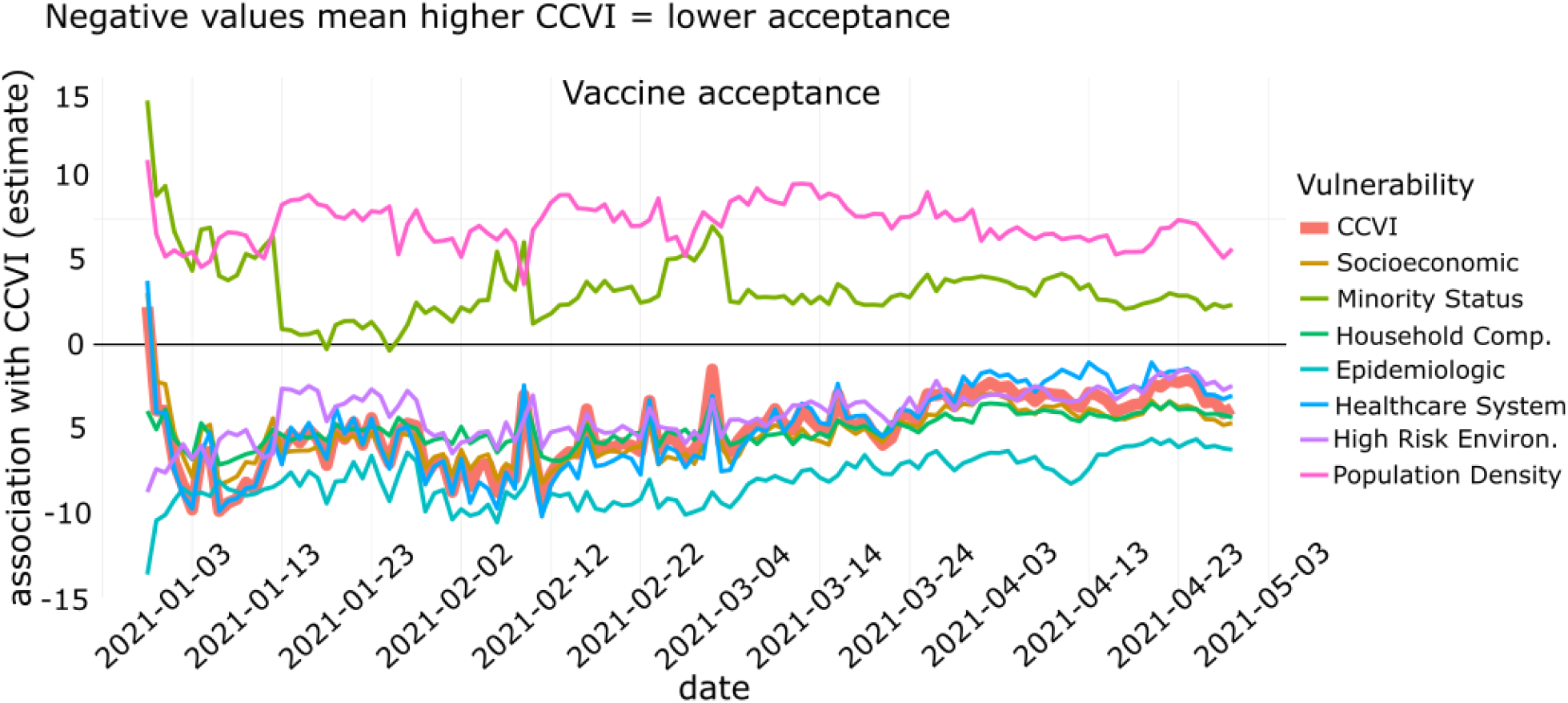
Regressions are simple univariate regressions, no mixed effects, weighted by county population. The dependent variable is the cumulative percentage of vaccine acceptors that have either already been vaccinated or definitely or probably would get the vaccine if offered to them. The coefficient of variance from each daily regression is plotted as a time series.

## Discussion

Our findings build on emerging evidence that vulnerable communities across the US have sustained the brunt of the negative impacts from the COVID-19 pandemic, and these adverse effects are being prolonged by lower vaccine uptake.^6,11–13^ The results show that more vulnerable counties have lower vaccine coverage, both in terms of first doses administered and the population fully vaccinated, compared to less vulnerable counties during the study period. This association became stronger throughout April into early May indicating that the gap in vaccination coverage is widening for vulnerable populations. The vulnerability dimensions that are most strongly associated with lower vaccination coverage include Housing Type & Household Composition, Healthcare System Factors, and Socioeconomic Status. We also found that denser urban settings tend to be positively associated with higher vaccination coverage resulting in rural locations to fall behind.

The CDC has warned that disparities in vaccination coverage between urban and rural environments has been widening, and our results support that claim.^4^ We have shown that vulnerability matters both among counties across states as well as within states meaning that state-level differences are not the only drivers of variability. We hypothesize that this trend would continue into individual neighborhoods and census tracts if we had the granular data to assess such localized levels. Several themes deviate in their associations with vaccination coverage. For instance, greater Epidemiologic Risk Factors are associated with higher coverage among counties within a state only. This is most likely a consequence of some states prioritizing populations with preexisting conditions when supplies were scarce resulting in these counties receiving greater coverage within that state.

We also observed more vulnerable counties associated with lower levels of vaccine acceptance. However, when considering the overall CCVI metric, the gap in vaccination coverage between high and low vulnerable counties cannot be explained solely by vaccine hesitancy sentiments. Lower vaccine acceptance rates did associate with higher vulnerability, and this could exacerbate some of the inequities in vaccination coverage. However, the percentage difference in vaccine acceptance between high and low vulnerable counties accounts for only about 6% of the difference in vaccination coverage in those same counties. This finding implies that there are systemic differences between high and low vulnerability counties driving the disparities in vaccination coverage, and these need to be addressed rapidly through policy interventions.

Many states and local jurisdictions have been using social and other disadvantaged vulnerabilities to prioritize geographies for vaccine uptake.^5^ However, we continue to see a wide gap in vaccination coverage among the vulnerable. Schmidt et al. 2021 report that as of March 30, 2021, only 37 jurisdictions, including 34 states, had adopted the use of disadvantage indices^6,14^ and place-based measures such as monitoring receipt to reduce inequity. To ensure that equity features centrally in allocation plans and to reduce the gap in coverage, policymakers should continue to focus on vulnerable communities through the use of vulnerability indices and targeted interventions.

## Conclusion

The COVID-19 pandemic has highlighted the inequities of US communities across a range of social, health, and economic vulnerabilities. Evidence shows that vulnerable communities have experienced far worse outcomes throughout the pandemic, and our findings of higher vulnerability counties having lower vaccination coverage suggest these negative impacts are continuing throughout the vaccine rollout. While lower vaccine acceptance is associated with higher vulnerability, the magnitude of hesitant sentiments cannot fully explain the disparities in coverage. This suggests various systemic issues need to be addressed to lower the gap for vulnerable communities. Vaccination coverage is affecting the disadvantaged across a range of equity dimensions. Therefore, the use of vulnerability indices that create a composite of these different dimensions can be useful in prioritizing and allocating additional aid. While many jurisdictions across the US have seen an increased use in vulnerability indices to guide policy, this needs to become universal before the vaccination gap can be closed.

## Limitations

The community-level design with county-level outcome metrics does not allow for inequities at the individual level to be investigated. The vaccine acceptance data coverage accounts for only ∼50% of US counties and ∼61% of the US population making the results slightly biased toward counties with larger populations. Due to data availability issues at the county-level, we only conducted the study for the month of April and early May. Therefore, the study is not representative of the entire vaccine rollout. While vulnerability is associated strongly with vaccination coverage, there may be other contributing factors to the vaccination gap that have not been analyzed in this study, such as state and local policies.

## Data Availability

The data are available in a public, open access repository. The COVID-19 Community Vulnerability Index and an interactive data explorer are available at the Surgo Venture website. It is made available under a CC-BY-NC 4.0 International license. The county-level vaccination rates are available through the Covid Act Now public API. The county-level vaccine acceptance data is available through the COVIDcast public API.

https://precisionforcovid.org/ccvi

https://covidactnow.org/data-api

https://delphi.cmu.edu/covidcast/indicator/?date=20210604&sensor=fb-survey-smoothed_covid_vaccinated_or_accept

## Data Availability

The data are available in a public, open access repository. The COVID-19 Community Vulnerability Index and an interactive data explorer are available at the Surgo Venture website. It is made available under a CC-BY-NC 4.0 International license. The county-level vaccination rates are available through Covid Act Now’s public API. The county-level vaccine acceptance data is available through COVIDcast’s public API.

https://precisionforcovid.org/ccvi

https://covidactnow.org/data-api

## Acknowledgements

We thank Covid Act Now for their work in providing longitudinal county level vaccination data. We also thank the Carnegie Mellon University Delphi Group for providing detailed county-level survey data on vaccine acceptance.

## References

1. Dong E, Du H, Gardner L. An interactive web-based dashboard to track COVID-19 in real time. The Lancet. 2020;20(5):533-534. doi:https://doi.org/10.1016/S1473-3099(20)30120-1

2. Kim SJ. Social Vulnerability and Racial Inequality in COVID-19 Deaths in Chicago. Health Educ Behav. 2020;47(4). doi:https://doi.org/10.1177/1090198120929677

3. Surgo Ventures. COVID-19 COMMUNITY VULNERABILITY INDEX. Precision for COVID. Published June 10, 2021. https://precisionforcovid.org/ccvi

4. Murthy BP, Sterrett N, Weller D, Zell E. Disparities in COVID-19 Vaccination Coverage Between Urban and Rural Counties — United States, December 14, 2020–April 10, 2021. CDC; 2021:759–764. http://dx.doi.org/10.15585/mmwr.mm7020e3externalicon

5. Schmidt H, Weintraub R, Williams MA, et al. Equitable allocation of COVID-19 vaccines in the United States. Published online May 18, 2021. doi:https://doi.org/10.1038/s41591-021-01379-6

6. Smittenaar P, Stewart N, Sutermaster S, et al. A COVID-19 Community Vulnerability Index to drive precision policy in the US. medRxiv. Published online May 20, 2021. doi:https://doi.org/10.1101/2021.05.19.21257455

7. Coivd Act Now. Covid Act Now Data API.; 2021. https://covidactnow.org/data-api

8. CMU Delphi. COVIDcast Epidata API. https://delphi.cmu.edu/covidcast/indicator/?date=20210604&sensor=fb-survey-smoothed_covid_vaccinated_or_accept

9. Raudenbush SW, Bryk AS. Hierarchical Linear Models: Applications and Data Analysis Methods. 2nd ed. Sage Publications, Inc.; 2002.

10. Snijders TA, Bosker RJ. Multilevel Analysis : An Introduction to Basic and Advanced Multilevel Modeling. 2nd ed. Sage Publications, Inc.; 2012. https://lib.ugent.be/catalog/rug01:001698339

11. Stokes EK, Zambrano LD, Anderson KN. Coronavirus Disease 2019 Case Surveillance — United States, January 22–May 30, 2020. CDC; 2020:759-765. DOI: http://dx.doi.org/10.15585/mmwr.mm6924e2externaliconexternalicon

12. Office of Disease Prevention and Health Promotion. Social Determinants of Health | Healthy People 2020. Published 2021. https://www.healthypeople.gov/2020/topics-objectives/topic/social-determinants-of-health

13. Kantamneni N. The impact of the COVID-19 pandemic on marginalized populations in the United States: A research agenda. J Vocat Behav. 2020;119. doi:doi:10.1016/j.jvb.2020.103439

14. Flanagan B, Gregory E, Hallisey E, Heitgerd J, Lewis B. A Social Vulnerability Index for Disaster Management. J Homel Secur Emerg Manag. 2011;8(1). doi:10.2202/1547-7355.1792

