## Supplemental Material for "Inequities among vulnerable communities during the COVID-19 vaccine rollout"

### Appendix 1

#### Average difference in Vaccination Coverage among CCVI Terciles on May 8

eTable 1. Average full vaccination coverage across counties in the top and bottom terciles of vulnerability as defined by the US COVID-19 Community Vulnerability Index.

| Dimension | Most vulnerable third of counties: full vaccination coverage | Least vulnerable third of counties: full vaccination coverage | % Difference: full vaccination coverage (least vs most vulnerable) |
|---|---|---|---|
| **CCVI** | 30.7% | 34.6% | +12.7% |
| **Socioeconomic** | 30.2% | 35.3% | +16.9% |
| **Minority Status & Language** | 32.7% | 30.9% | -5.5% |
| **Housing Type & HH Comp.** | 27.6% | 34.8% | +26.1% |
| **Epidemiological** | 29.6% | 33.2% | +12.2% |
| **Healthcare System** | 31% | 34.1% | +10% |
| **High Risk Environments** | 30.9% | 33.1% | +7.1% |
| **Population Density** | 32.9% | 27.7% | -15.8% |

### Average difference in Vaccine Acceptance among CCVI Terciles on May 8

eTable 2. Average vaccine acceptance across counties in the top and bottom terciles of vulnerability as defined by the US COVID-19 Community Vulnerability Index.

| **Dimension** | **Most vulnerable third of counties: vaccine acceptance coverage** | **Least vulnerable third of counties: vaccine acceptance coverage** | **% Difference: vaccine acceptance coverage (least vs most vulnerable)** |
|---|---|---|---|
| **CCVI** | 85.5% | 86.1% | +0.7% |
| **Socioeconomic** | 85.95% | 87.3% | +1.6% |
| **Minority Status & Language** | 87.5% | 80% | -8.6% |
| **Housing Type & HH Comp.** | 82.6% | 87.2% | +5.6% |
| **Epidemiological** | 80.2% | 87.8% | +9.5% |
| **Healthcare System** | 86.1% | 86% | +0.1% |
| **High Risk Environments** | 82.9% | 87.7% | +5.8% |
| **Population Density** | 86.9% | 79.3% | -8.8% |
